# Proposing Response Evaluation Criteria in Solid Tumors Based on Genomic Profiling or Genomic RECIST: A Retrospective Study on the Liquid Biopsy Results of 29 Cancer Patients

**DOI:** 10.1101/2021.12.13.21266491

**Authors:** Khin Zay Yar Myint, Masamori Shimabuku, Kenichi Kashihara, Ruriko Horio, Munehisa Kaneda, Yoko Shimizu, Junichi Taguchi

**Author notes:** Corresponding author: Junichi Taguchi, MD, PhD, Director, Center of Advanced Medical Science and Technology, Tokyo Midtown Medical Center, 9-7-1, Akasaka, Minato-ku, Tokyo, 107-6206.

## Abstract

Tumor response and disease progression are assessed using imaging technologies. However, these technologies fail to detect tumor responses at the molecular level and clonal evolution. A potential surrogate for such parameters is using circulating tumor DNA (ctDNA). This study aimed to examine the quantity and composition of the ctDNA results of 29 cancer patients before and after dendritic cell (DC) immunotherapy and develop criteria to evaluate the molecular response to treatment based on these results. We categorized the patients into four categories based on percent changes in the total ctDNA compared with the baseline ctDNA titers, and this response assessment was termed genomic response evaluation criteria in solid tumors or gRECIST. Even those who are clinically evaluated as having a good response might harbor unfavorable tumor responses at the molecular level. Newly formed ctDNA levels can be the most prognostic parameter in tumor progression or the treatment response, while ctDNA clearance and the decline or rise in existing ctDNA did not change significantly in genomic response categories (gRECIST). More research is needed to support the clinical use of ctDNA in precision oncology and personalized cancer treatment.

## Introduction

Overall survival is the most preferable end of therapeutic efficacy in cancer research. However, the tumor response to treatment and time to disease progression—i.e., changes in the tumor burden—are also important to clinically evaluate the effectiveness of certain cancer treatments in clinical trials. These changes are usually assessed anatomically using radiological images. The first standardized assessment tool was developed by the World Health Organization (WHO) in 1981. It assessed tumor burden using the sum of the products of diameters (SPDs) and determined the response to therapy by evaluating changes from baseline during treatment [1]. Some common criticisms were raised against the WHO criteria such as the following: using SPD can underestimate the actual response rate if a minor change in the tumor size or a measurement error occurs, and no clear definitions exist for the number and minimum size of lesions to be measured or disease progression [2]. As the criteria have been modified in clinical trials to address these unclear points, confusion emerged over the interpretation of the results, and it became challenging to make conclusions about the efficacy of the same regimen [3-4].

In the mid-1990s, an International Working Party tried to establish simple and standardized guidelines for the evaluation of therapeutic efficacy. These guidelines were released in 2000 [5] and called Response Evaluation Criteria in Solid Tumors (RECIST). The original RECIST criteria (RECIST version 1.0) addressed the above criticisms of the WHO criteria. Key features include the use of unidimensional (i.e., the longest diameter) measures of the tumor, instructions on the number of lesions to follow, definitions of the minimum size of measurable lesions, and progressive disease. It also included guidelines on using new technology at that time, such as spiral CT. Since then, the RECIST criteria have become widely used and accepted assessment criteria to evaluate the therapeutic efficacy of cancer treatment, particularly new anticancer agents, in clinical trials. It was revised to RECIST version 1.1 in 2009 based on the analysis of more than 6500 patients [6-7]. The new version included additional features such as the assessment of lymph nodes and guidance on multidetector CT and magnetic resonance (MR) imaging.

Both WHO and RECIST used criteria based on anatomic size and lesion changes during treatment using radiological technologies. These changes were categorized into four groups: (a) complete response, (b) partial response, (c) stable disease, and (d) progressive disease. Therefore, these tools mostly assess gross tumor shrinkage only as an indicator of the treatment response or tumor progression but no other antitumor activities of the given treatment. Furthermore, these criteria were primarily developed to evaluate the therapeutic efficacy of cytotoxic drugs. As cancer treatment has advanced, become more targeted and personalized in recent years, using RECIST alone is challenging to assess the efficacy of other treatment methods, such as molecular-targeted therapies, locoregional therapies or immunotherapies. Additionally, it takes time (several weeks or sometimes months from initiation of treatment) to evaluate the treatment response using imaging technology. Treatment response should be continuously and timely monitored so that appropriate and prompt actions can be taken in accordance with the therapeutic results.

Such a gap can be filled using genomic profiling technology of liquid biopsies. Liquid biopsy, which is the analysis of free circulating tumor DNA (ctDNA) from blood or plasma, can noninvasively monitor the tumor burden within a short time. Circulating tumor DNA is cancer-derived DNA resulting from the apoptosis or necrosis of tumor cells. It is a short fragment of DNA of approximately 134-144 base pairs [8]. It can reflect the response of the whole tumor mass, not a part of the tumor, as well as real-time genomic alterations, which are critical in detecting clonal evolution. Thus, it can help detect emerging resistant mutations and mutated DNA from different metastatic sites. It can also be performed repeatedly and serially to monitor disease progression at appropriate intervals. Several studies have proven that liquid biopsy can be a time-saving, less-invasive, and up-to-the-minute analysis that could provide valuable insights into the treatment response and tumor progression [9-11].

In this study, we aimed to examine the quantity and composition of ctDNA results of 29 cancer patients before and after undergoing dendritic cell (DC) immunotherapy with or without chemotherapy and/or radiotherapy and developed criteria to evaluate the molecular response to treatment based on these results.

## Materials and methods

### Study population

This was a retrospective observational study of genomic profiling results in 29 cancer patients who had undergone dendritic cell immunotherapy at the Department of Advanced Medical Science and Technology, Tokyo Midtown Medical Center. Blood specimens for liquid biopsy were taken from the patients immediately before and after completion of one course of DC immunotherapy.

### Ethical approval and informed consent

The study was carried out in accordance with the Declaration of Helsinki and approved by the Institutional Review Board of Midtown Clinic Medical Corporation. We obtained written informed consent before the blood tests, after explanation of the nature of the blood test, use of their data for clinical research, and their right to withdraw anytime.

### Genomic profiling using liquid biopsy

We used GenoDive (GenoDive Pharma, Atsugi, Kanagawa, Japan) assays for genomic profiling of cancer patients in this study. The GenoDive assays were performed at the GenoDive Pharma Okinawa Branch. Tumor profiling was performed at the laboratory using the AVENIO ctDNA Expanded Kit (Roche Sequencing Solutions, Pleasanton, CA). All the methods and procedures were performed in accordance with relevant guidelines and regulations. AVENIO ctDNA kits were reported as reliable tools for detecting somatic mutations in clinical laboratories [12, 13].

### Dendritic cell (DC) immunotherapy

#### Preparation of dendritic cell (DC) vaccines

DC vaccines were prepared in accordance with the standard operation procedures and quality control measures as reported previously [14-17]. First, we collected peripheral blood mononuclear cells via leukapheresis using a specialized machine (Spectra Optia®, Terumo BCT, Inc., Tokyo, Japan). This machine isolates only the required white blood cells by centrifugation and returns the other blood components, such as red blood cells, platelets, plasma, and other white blood cells, such as neutrophils, back into the circulation so that total blood loss could be minimal. Monocytes were isolated from white blood cells and then cultured with human recombinant granulocyte macrophage colony-stimulating factor (GM-CSF) and human recombinant interleukin-4 (IL-4) to transform them into immature DCs. On the sixth day of culture, the DCs were activated with OK-432 and prostaglandin E2 for 24 hours. At the same time, the DCs were pulsed with two types of artificial cancer antigens, Wilms tumor gene 1 (WT1) and Mucin 1, cell surface associated (MUC1) antigens. The DCs were then cryopreserved until the date of administration.

#### Administration of DC vaccines

DC vaccines were administered intradermally to areas close to lymph nodes such as the groin, axilla and/or neck every two to three weeks for at least 7 times (one course).

#### Safety and quality

High-quality GM-CSF and IL-4 were used to culture dendritic cells at our center to avoid unfavorable incidents and ensure patient safety. When the culture was complete, the dendritic cells were washed thoroughly to limit contamination of reagents. Additionally, the cells were assessed for the absence of harmful agents such as bacteria, fungi, viruses or endotoxins. Specific infection control measures were adopted for patients with infectious diseases. Dendritic cell immunotherapy is generally a safe procedure to be performed on an outpatient basis. No serious adverse events were reported in our study cohort.

### Setting genomic RECIST criteria

Using the extent of tumor shrinkage in RECIST 1.1 criteria as cutoff values, Dr. Taguchi, a clinical geneticist, categorized ctDNA changes into four categories, and this response assessment was termed genomic RECIST. The cutoff values for genomic RECIST and reference values from RECIST 1.1 criteria are shown in Table 2. The change in total ctDNA was calculated as the percent of total ctDNA after the treatment (LB2) as the numerator and the percent of total ctDNA at baseline (LB1) as the denominator. The percentage of total ctDNA was the titer of ctDNA divided by that of cell-free DNA (cfDNA). The baseline ctDNA percent (LB1) must be at least 0.1% to be eligible for this calculation. In other words, we set the limit for the denominator at 0.1 to avoid extremely large and infinite values for ratios in total ctDNA (LB2/LB1). In one case, LB1 was zero, and it was excluded from the analysis. The first category is gCR, in which the LB2/LB1 ratio was set as less than 0.01. This cutoff value was referenced from the clinical RECIST criteria for CR—that is, the disappearance of all target lesions and any pathological lymph nodes less than 10 mm reduction in the short axis. Because clinical RECIST criteria for PR require at least a 30% decrease in the sum of diameters of target lesions, considering the baseline sum diameters as a reference, the cutoff values of LB2/LB1 for gPR were set between 1% and 70%. Clinically, progressive disease was defined as a tumor size with a 20% relative increase from baseline; therefore, the cutoff value of LB2/LB1 for gPD was set as above 120%. Finally, the values of LB2/LB1 between gPR and gPD, which are between 70% and 120%, were categorized as genomically stable disease or gSD. Next, we calculated the percent clearance, decline, newly formed and increase in ctDNA using LB1 as a denominator.

### Changes in ctDNA regarding treatment regimens and the clinical response

Changes in the quantity and composition of circulating tumor DNA (ctDNA) levels were first compared among patients who had received different treatment regimens while undergoing DC immunotherapy. These changes were also compared among those diagnosed with different clinical stages after dendritic cell immunotherapy.

### Statistical methods

All continuous variables were expressed as means and standard deviations. All categorical variables were expressed as numbers and proportions. The calculations and figure generation were performed using Microsoft Excel (Microsoft Corporation, 2018) and R software (R 4.1.0, R Core Team, 2021).

## Results

The demographic, clinical and pathological characteristics of the study cohort are summarized in Table 1. The study cohort comprised 29 patients, aged 35 to 91 years. More than half of the patients were seniors in their 60 s to 70 s. The sex of the population was also balanced in this cohort. Almost all the patients were diagnosed with cancer for the first time, and only two of them had recurrence of the primary tumor. Approximately 80% of them were diagnosed at an advanced stage of cancer, and more than 80% of them had metastasis to at least one site outside the primary tumor. Only two of them had no prior treatment before DC immunotherapy. Surgery with adjuvant chemotherapy was the most common treatment combination (n=12), followed by chemotherapy (n=6), surgery (n=4), all conventional therapies (n=3), and radiotherapy and chemotherapy (n=2). Regarding treatment regimens during DC immunotherapy, 18 patients received combination chemotherapy, 1 patient received radiotherapy, and 3 patients received both chemo- and radiotherapy. Only 7 of the patients received DC immunotherapy alone. The duration and dosage of DC immunotherapy varied among patients. The average interval between two liquid biopsy tests was 116 days (range: 76 to 232 days). The average dosage of live cells was 81.01% (range: 59.74% to 88.2%) per ten million dendritic cells. The tumor types included in this study cohort were pancreatic cancer (n=9), colorectal cancer (n=4), stomach cancer (n=3), non-small cell lung cancer (NSCLC, n=2), breast cancer (n=2), and biliary cancer (n=2), and the remaining cancer types were peritoneal carcinomatosis, sarcoma, Paget’s disease, uterine cancer, pancreatic neuroendocrine tumor, apocrine gland cancer, and ureteral cancer (Figure 1).

**Table 1:** Summary of demographic, clinical and pathological characteristics of the study cohort.

| Characteristics | Participants<br>(n=29) |  |
| --- | --- | --- |
| Age, average (range) | 67 (35 – 91) |  |
| Sex, n (%) |  |  |
| Male | 15 | (52%) |
| Female | 14 | (48%) |
| Diagnosis, n (%) |  |  |
| First time | 27 | (93%) |
| Recurrence | 2 | (7%) |
| Metastases, n (%) |  |  |
| No metastasis | 5 | (17%) |
| One site | 6 | (21%) |
| Multiple sites | 18 | (62%) |
| Clinical staging, n (%) |  |  |
| I | 1 | (3%) |
| II | 4 | (14%) |
| III | 1 | (3%) |
| IV | 23 | (79%) |
| Conventional treatment before DC, n (%) |  |  |
| Nil | 2 | (7%) |
| Surgery only | 4 | (14%) |
| Chemotherapy only | 6 | (21%) |
| Surgery and chemotherapy | 12 | (41%) |
| Radiotherapy and chemotherapy | 2 | (7%) |
| Surgery, chemotherapy and radiotherapy | 3 | (10%) |
| DC treatment regime, n (%) |  |  |
| DC only | 7 | (24%) |
| DC with chemotherapy | 18 | (62%) |
| DC with radiotherapy | 1 | (3%) |
| DC with both radiotherapy and chemotherapy | 3 | (10%) |
| Treatment interval in days, mean (range) | 116 | (76-232) |
| DC dosage or percent of live cells in ten million dendritic cells, mean (range) | 81.01 | (59.74-88.2) |

**Figure 1.**
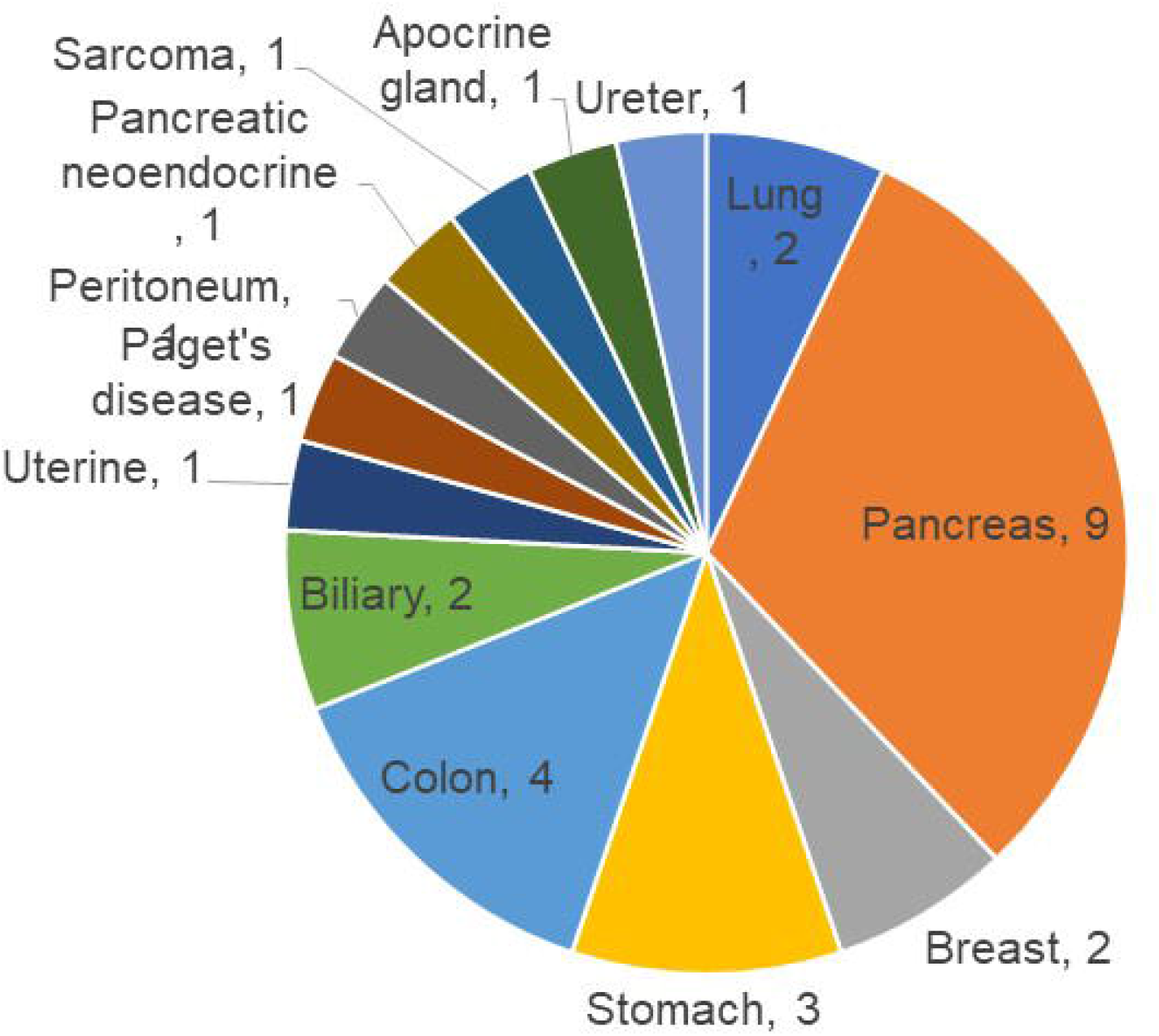
Types of cancer in the study cohort. Gastroenterological cancers such as pancreatic, colorectal, stomach, and biliary cancers constituted more than half of the population in the study cohort.

Table 2 shows the criteria for genomic RECIST based on the percent changes in total ctDNA before and after DC therapy (%change in total ctDNA) and respective numbers of patients in each category. No patient was categorized as having a genomic complete response (gCR), whose change in total ctDNA was less than 0.01 or 1%. Seven patients with a % change in total ctDNA greater than or equal to 1% and less than 0.7 or 70% were categorized as having a genomic partial response (gPR). Eight patients were categorized as having genomic stable disease (gSD), in which the % change in total ctDNA was greater than or equal to 0.7 or 70% and less than or equal to 1.2 or 120%. Finally, 14 patients were categorized as having genomic progressive disease (gPD), in which the % change in total ctDNA was above 1.2 or 120%.

**Table 2:** Proposed criteria for genomic RECIST.

| Category | Total ctDNA changes* (LB2/LB1 ratio) | Number of patients (n) | RECIST 1.1 tumor response criteria as a reference for cutoff values |
| --- | --- | --- | --- |
| gCR | <0.01 | 0 | Complete Response (CR): Disappearance of all target lesions. Any pathological lymph nodes (whether target or nontarget) must have reduction in short axis to <10 mm. |
| gPR | 0.01-0.69 | 7 | Partial Response (PR): At least a 30% decrease in the sum of diameters of target lesions, taking as reference the baseline sum diameters. |
| gSD | 0.7-1.2 | 8 | Stable Disease (SD): Neither sufficient shrinkage to qualify for PR nor sufficient increase to qualify for PD, taking as reference the smallest sum diameters while on study. |
| gPD | >1.2 | 14 | Progressive Disease (PD): At least a 20% increase in the sum of diameters of target lesions, taking as reference the smallest sum on study (this includes the baseline sum if that is the smallest on study). In addition to the relative increase of 20%, the sum must also demonstrate an absolute increase of at least 5 mm. |

\* Calculation: The change in total ctDNA was calculated as the percent of total ctDNA after the treatment (LB2) as the numerator and the percent of total ctDNA at baseline (LB1) as the denominator. The percentage of total ctDNA was the titer of ctDNA divided by that of cell-free DNA (cfDNA). Baseline ctDNA must be at least 0.1% to be eligible for this calculation.

Figure 2 shows the comparison of the number of patients in each clinical response and genomic response criterion. Clinical responses were evaluated using RECIST version 1.1. One patient was diagnosed as having a complete response, 6 as having partial responses, 2 as having between a partial response and stable disease, 16 as having stable disease, 1 as having between stable disease and progressive disease, and 3 as having progressive disease. Clinically, only 4 patients were diagnosed with progressive disease (PD), but 14 patients in the progressive disease category were identified by genomic RECIST.

**Figure 2.**
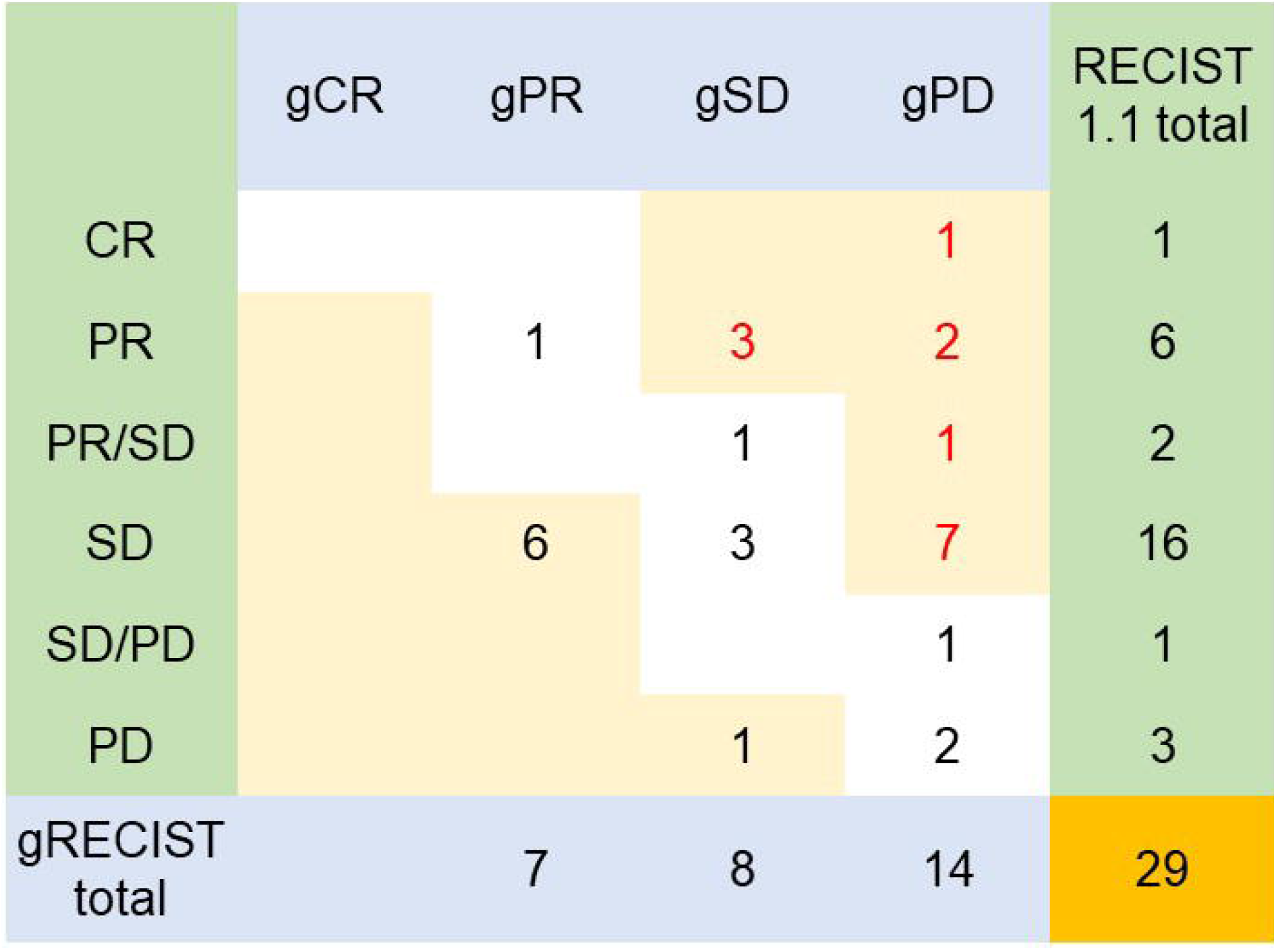
Comparison of clinical response and genomic response criteria. Even those who are clinically evaluated as having a good response might harbor unfavorable tumor responses at the molecular level.

Figure 3 shows the clonal evolution results in each genomic RECIST category. The values were calculated using total ctDNA at baseline as a denominator. Even in patients with gPR, newly formed ctDNA constituted approximately 30% of the total ctDNA. In patients with gSD, the percentages of clearance and new formation of ctDNA were almost the same, constituting approximately 40% of total ctDNA. In cases of gPD, the percentage increase in newly formed ctDNA was significantly high, whereas the existing ctDNA did not increase significantly (p=0.025). In trend analysis using analysis of variance (ANOVA), these trends were statistically significant at p<0.05.

**Figure 3.**
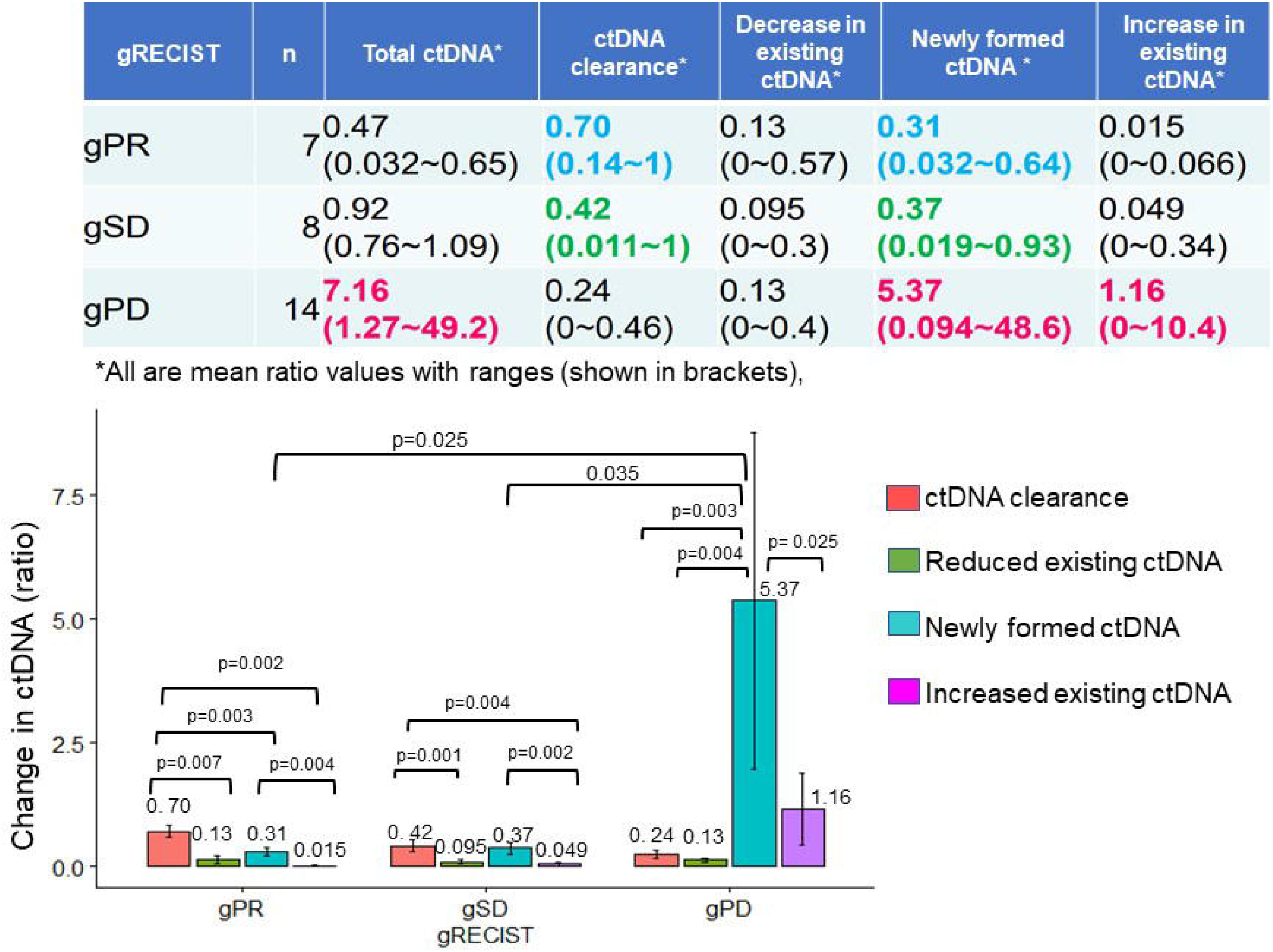
Clonal evolution results in each genomic RECISTcategories. Newly formed ctDNA levels can be the most prognostic parameter in tumor progression or treatment response, while ctDNA clearance and the decline or rise in existing ctDNA did not change significantly in genomic response categories (gRECIST).

Figure 4 shows the changes in ctDNA among patients who had received different treatment regimens while undergoing DC immunotherapy. The percent changes in total ctDNA and newly formed ctDNA were the most striking in those who had undergone both chemotherapy and radiotherapy during DC immunotherapy. Patients who had undergone DC immunotherapy alone had the least decrease in ctDNA compared with those with a combination of chemotherapy or both chemotherapy and radiotherapy. Figure 5 shows the comparison of percent changes in ctDNA among those diagnosed with different clinical stages after dendritic cell immunotherapy. We observed that even in clinically stable cases, high ctDNA due to newly formed ctDNA could occur, although the existing ctDNA level decreased. A significantly increasing trend in existing ctDNA was observed as the diseases progressed clinically.

**Figure 4.**
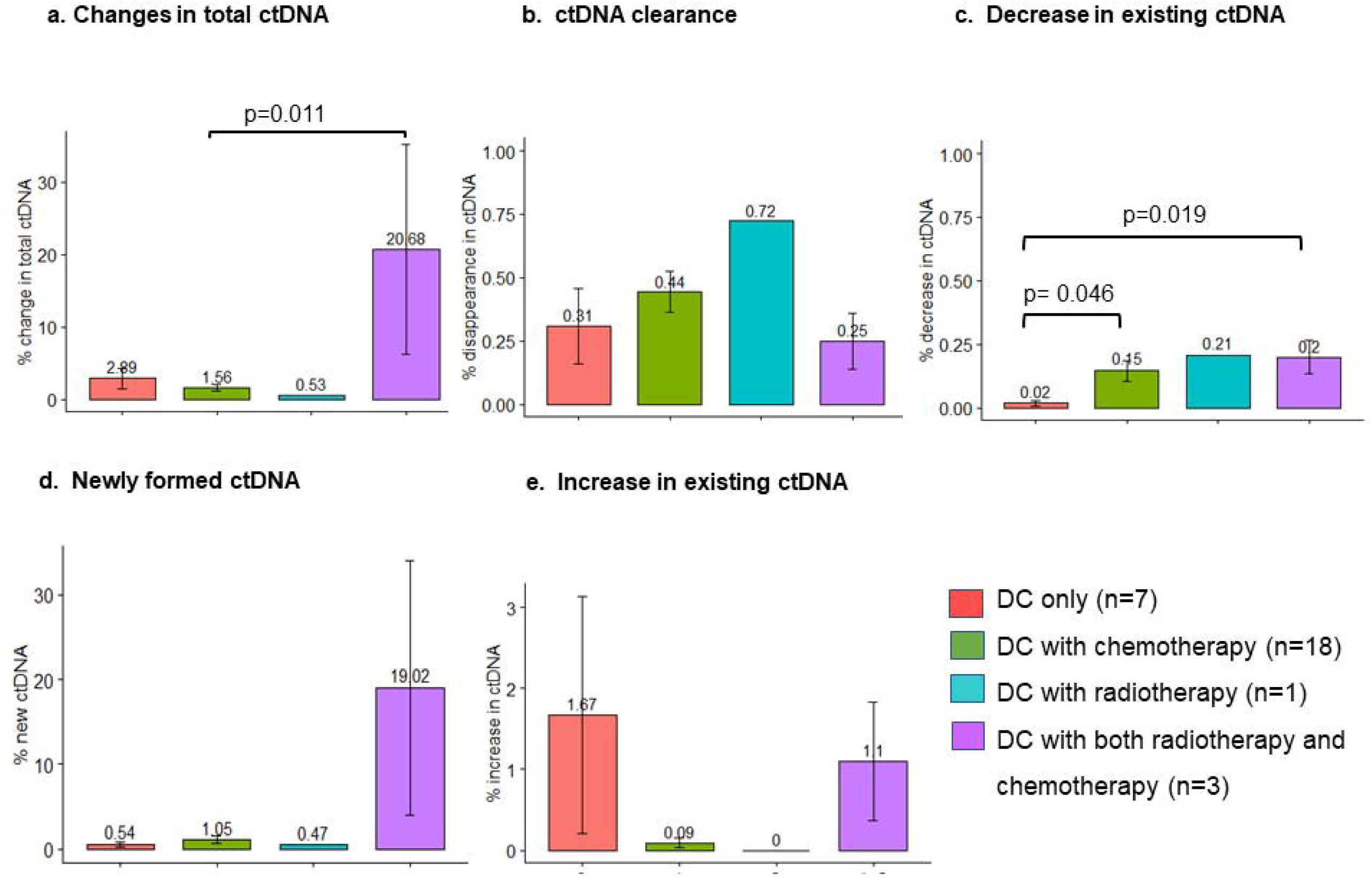
Changes in ctDNA with regards to treatment regimens. The patients who received both chemotherapy and radiotherapy during DC immunotherapy had the highest percent changes in total ctDNA and newly formed ctDNA. Patients who underwent DC immunotherapy alone had the least decrease in ctDNA.

**Figure 5.**
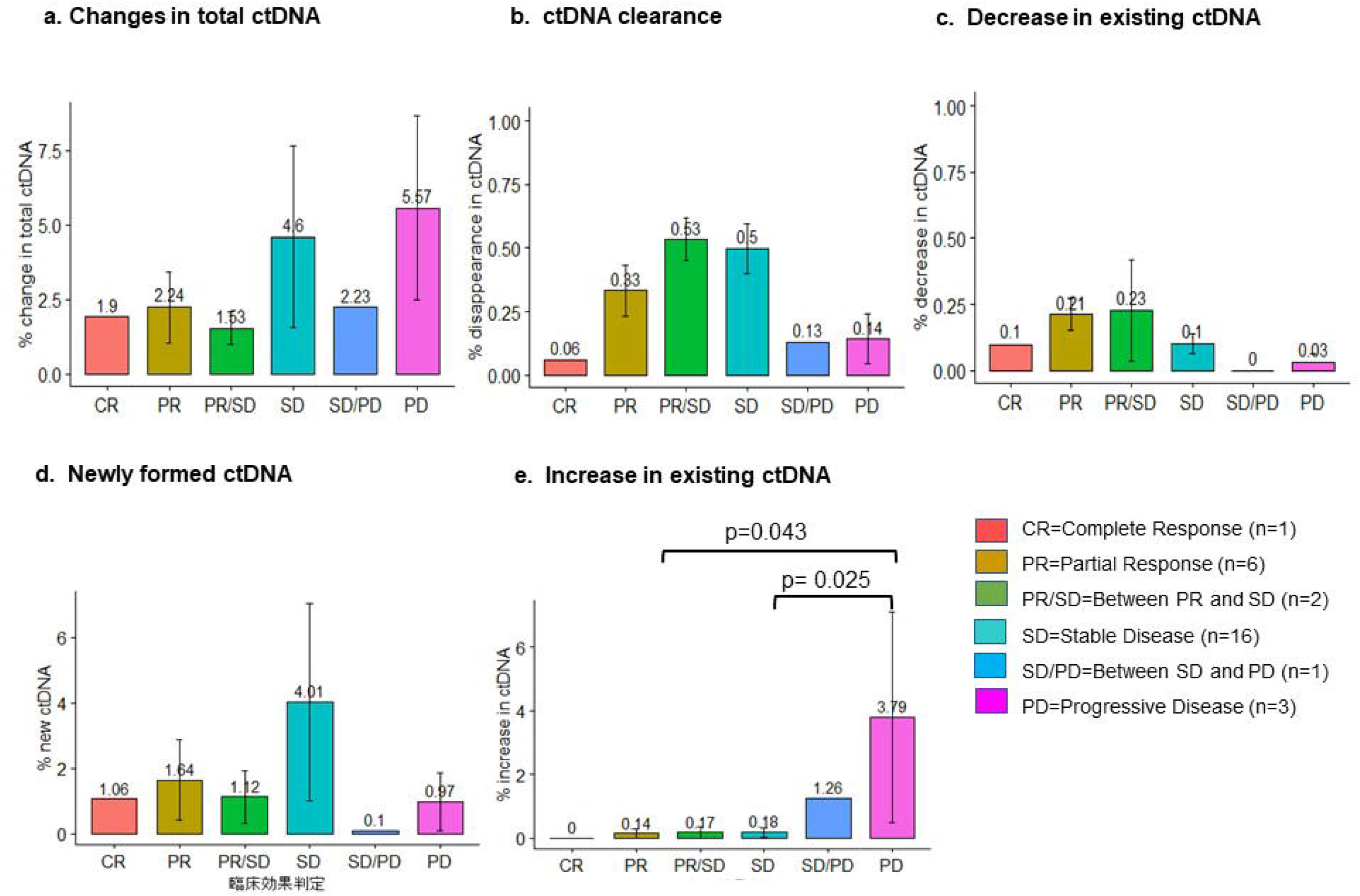
Changes in ctDNA with regards to clinical response. Even in clinically stable cases, there could be high ctDNA due to newly formed ctDNA. A significantly increasing trend in existing ctDNA was observed as the diseases progressed clinically.

## Discussion

Responses to treatment and disease progression are usually evaluated using imaging technologies. However, they cannot evaluate tumor responses at the molecular level or clonal evolution in response to treatment or the natural course of disease processes. A potential surrogate for such a parameter is using circulating tumor DNA (ctDNA), which is a prognostic marker in precision oncology and personalized cancer treatment. Its quantity and composition correlate with tumor stages and disease progression, such as recurrence, resistance and metastasis [18-20]. However, it has limited clinical benefit because it still lacks the requirements for integration in the clinical routine, such as analytical validity, reliability, and reproducibility [21]. Therefore, more practical studies are necessary in a real clinical setting to widen its clinical benefit. In the present study, we proposed the genomic RECIST criteria based on changes in the amount of ctDNA in the liquid biopsies of 29 patients before and after undergoing DC immunotherapy with or without chemotherapy/radiotherapy.

Most of the cases in this study cohort were patients at advanced stages of cancer with distant metastasis. Gastroenterological cancers such as pancreatic, colorectal, stomach, and biliary cancers constituted more than half of the population in the study cohort. Almost all of them, except two patients, had undergone at least one type of standard cancer treatment before undergoing DC immunotherapy. Only two of the patients had no prior treatment before DC immunotherapy. Except those patients, the remaining patients had undergone at least one type of conventional cancer treatment before DC immunotherapy, and surgery followed by chemotherapy was the most common treatment combination. Chemotherapy was the most common treatment combined with DC immunotherapy in this study cohort.

Song et al. reported that ctDNA clearance was correlated with the treatment response and disease progression in a multicenter prospective study of 949 advanced NSCLC patients [10]. In our study, we found that not only ctDNA clearance but also newly formed ctDNA levels can be prognostic in tumor progression or treatment response. In those categorized as gPR, approximately 30% of newly formed ctDNA was possible, reflecting the dynamic state of tumor evolution. However, a partial response at the molecular level was assessed because the percent ctDNA clearance surpassed that of newly formed ctDNA, and the percent decrease surpassed the percent increase in existing ctDNA. In gSD, the clearance and new formation were balanced, as well as the percent decrease and increase in existing ctDNA. The most striking characteristic of gPD was the significant formation of new ctDNA, while ctDNA clearance and the decline or rise in existing ctDNA did not change significantly compared with other categories.

Discrepancies existed between clinical and genomic responses, particularly those evaluated as having a good response clinically that might harbor unfavorable tumor responses at the molecular level. For example, even in clinically stable cases, ctDNA can be high because of newly formed ctDNA, although the existing ctDNA level decreases. Therefore, we can postulate that newly formed ctDNA values and compositions may be responsible for tumor progression. In other words, the prognosis of those cases might not be as good as clinically evaluated, and close monitoring of ctDNA values might be required to capture a more precise picture of the tumor response. Patel et al. reported that higher ctDNA levels were associated with a poor prognosis in pancreatic cancer patients [22]. Hence, monitoring ctDNA titers prospectively is critical in cancer prognosis and should be incorporated in the clinical monitoring of cancer patients.

The benefits of precision oncology using ctDNA should be extended not only to those after receiving standard cancer treatments to monitor the treatment response, prognosis and recurrence but also to those who are ineligible for standard treatments to guide the selection of appropriate treatment options. Cancer patients might have at least one type of potentially actionable genomic alteration [23]. However, a large gap exists between this genomic information and the availability of molecular-targeted therapy [24]. These valuable genomic profiling results must be applied to other emerging treatment options, such as immunotherapy. For example, personalized dendritic cell immunotherapy can be developed targeting major genomic alterations identified in the baseline liquid biopsy results of individual patients. The compositions of these alterations can be monitored to evaluate the treatment response in subsequent liquid biopsy tests. Serial blood testing can also track clonal changes, which can further guide the selection of potentially effective therapies. Our study has a few limitations. The first and most crucial limitation is the generalization of the findings because it was a small, single-center study in a specific patient population with varied clinical backgrounds. The type, dosage and duration of the treatment were also different. However, as an individual personalized assessment of cancer treatment, this study indicates the importance of using genomic response criteria to monitor treatment efficacy and tumor progression and provides insights for appropriate treatment selection.

## Conclusion

To our best knowledge, this study is the first and only to propose using genomic RECIST and liquid biopsy results in various tumors after DC immunotherapy with standard cancer treatments. Our study showed that genomic RECIST could be useful to monitor the treatment response, disease progression, and the selection of potential effective treatment in cancer patients. Additionally, our study highlighted that more delicate and precise assessment is feasible by prospectively monitoring ctDNA values. Although generalizability remains an issue, these findings have opened up further potential for real-world data and evidence to support the clinical use of ctDNA in precision oncology and personalized cancer treatment.

## Data Availability

All data produced in the present study are available upon reasonable request to the authors

## Acknowledgments

We would like to acknowledge Ms. Kumi Kimura and Ms. Hanano Sakurai for compilation of the relevant data for this study.

## Author contributions

KZYM was involved in data analysis and interpretation and drafted the manuscript.

JT is the principal investigator and corresponding author, who conceptualized or designed the entire study, conducted data analysis and revised the manuscript.

MS is the coinvestigator, who contributed to the conceptualization and implementation of the study and revision of the manuscript.

KK, RH, MK are the coinvestigators of the study.

YS contributed to the design and implementation of the study.

## Additional Information (including a Competing Interests Statement)

The authors declared no conflict of interest.

